# A Census of Clinical Trials Conducted Under the US Exception from Informed Consent Rule

**DOI:** 10.1101/2022.08.23.22279138

**Authors:** Krista L. Snyder, Jon F. Merz

## Abstract

**Background:** The US Food and Drug Administration and National Institutes of Health adopted the Exception from Informed Consent (EFIC) rule in 1996, permitting waiver of informed consent for certain emergency research, including trials funded by the federal government. The rule requires that prospective consent be sought when practicable from patients or their Legally Authorized Representative(s) (LAR), and for those enrolled without consent, the patient or their LAR must be given information and an opportunity to opt-out from continued participation at the earliest opportunity. We sought to census the trials conducted under the EFIC rule to facilitate research to better understand how the rule is being used.

**Methods:** We conducted a multi-pronged search to try and identify all trials conducted under the EFIC rule, drawing on numerous reviews, Medline and Google searches (including of the clinicaltrials.gov registry), examination of the FDA’s docket, posting an inquiry on the IRB Forum, and email requests to lead authors of all published EFIC trials and related review articles. We describe the trials, when they were started and completed, and whether they were terminated early.

**Results:** We identified a total of 105 trials as of April 1, 2022: 77 complete, 10 recruiting, 10 registered on clinicaltrials.gov but not yet recruiting, 5 trials that were abandoned before enrolling any subjects, and 3 trials in early planning. Nine of the 77 completed trials were pilot or feasibility trials. Of 68 completed full trials, 30 (44.1%) were terminated early. The most common reason for early termination was futility or safety (17 trials, 25.0%) followed by poor recruitment (9 trials, 13.2%). The rate of conduct of trials has been remarkably constant since 2001, with roughly 18 trials started in each 5-year period.

**Conclusions:** The rate of early termination of EFIC trials for futility or safety appears higher than for other kinds of clinical research. We provide the list of trials in a Supplement for further in-depth data collection and analysis of this set of trials.

---

Federal regulations in the United States have allowed research conducted under the auspices of the Food and Drug Administration (FDA) to be conducted without consent in emergency, life-saving situations since 1963.^1,2^ In the early 1990s, with increasing attention to the need to conduct such trials, the National Institutes of Health (NIH) were blocked from funding emergency research because such research was deemed to pose more than minimal risk to subjects.^3,4^ Minimal risk is a condition precedent for waiving consent under the Common Rule, which applies to all research funded by the federal government other than FDA.^5^ In 1996, the FDA promulgated a new, more detailed rule, the Exception from Informed Consent (EFIC).^6^ The EFIC rule was adopted by the NIH, permitting federal funding of emergency trials using the new waiver mechanism.

There have been 3 published reviews focused at least in part on EFIC trials. Klein, Moore & Biros performed a systematic review examining emergency trials conducted using waivers of consent (including the EFIC mechanism) in the 20 years since the rule was adopted.^7^ They identified 24 completed EFIC trials, 10 of which had been terminated early. Feldman, Hey & Kesselheim accessed the FDA’s docket 95S-0158 to identify all trials submitted for FDA approval between November 1, 1996 and October 23, 2017.^8^ They identified 41 trials, 37 of which had completed, at least 9 of which had terminated early (4 trials had no available data).

Armstrong and colleagues performed a systematic review examining consent practices for prehospital ambulance RCTs. They identified a total of 45 trials (4 of which were performed in the United States) conducted between 2000 and 2016.^9^ In addition, Haggins and colleagues reviewed the FDA docket, focusing on community consultation and public disclosures for proposed EFIC trials submitted through June, 2017. They identified 34 trials.^10^ Most recently, Neal Dickert and colleagues performed a scoping review, identifying 27 articles reporting results of community engagement activities for planned EFIC trials.^11^

We noted that there were some inconsistencies between reports, including trials we identified in a review performed by one of us.^12^ Thus, we sought to develop a census of EFIC trials to enable continued study, and to gain greater insights into the use of the EFIC mechanism. Here, we present only summary information on the process and the trials found in order to solicit information about any trials missed in this review, and to offer the census to others for further study.

## METHODS

We started with the trial samples identified by Klein et al.,^7^ Feldman et al.,^8^ Armstrong et al.,^9^ and Dhamanaskar & Merz.^12^ We then examined the papers in the reviews of community engagement activities conducted in support of EFIC trials by Haggins et al.,^10^ and Dickert et al.^11^ as an indirect way of identifying trials. Other articles and reviews were examined to ensure we were capturing proposed and completed EFIC trials.^13-17^ From December 2021 through April 2022, we searched PubMed and clinicaltrials.gov for “EFIC,” “50.24,” “exception”, “exception from informed consent” and combinations of these terms. We used Google Scholar for a general search for these same terms, and then used Google to specifically search the clinicaltrials.gov web page. We identified communicating authors of published papers and responsible investigators on clinicaltrials.gov, and emailed authors and investigators, clarifying ambiguous papers and asking if they knew of other trials. For papers focused on community engagement, we attempted to contact authors when necessary to identify the trials for which the community consultation was performed. We also posted a single inquiry on Public Responsibility in Research & Medicine’s (PRIMR) IRB Forum,^18^ asking for citations of any EFIC trials IRBs had reviewed. Finally, we searched the FDA’s public docket for submissions, primarily to double check our list of trials.^19^

We collected information about the trials, including disease or condition treated, whether the trial was a pilot or feasibility study, whether it focused on adults or children, years in which recruitment started and stopped, enrollments, and information about early termination and reasons for it. We planned descriptive statistics to examine patterns of recruitment.

Soliciting information about trials is not considered human subjects research by the University of Pennsylvania IRB.

## RESULTS

Fifty-two trials were identified from the 4 primary review articles.^7,8,9,12^ Our searches identified 37 more trials. Simultaneously, we emailed 99 authors or investigators, and received responses from 65 (65.6%). Respondents provided information on an additional 13 trials, as well as confirmed or clarified our search results. Our posting on the IRB Forum yielded one reply (off-list). Finally, tracking down trials from reviews focused on community engagement yielded 3 more trials. We note that our use of Google to search clinicaltrials.gov was highly effective, because it searched documents (including consent forms and protocols) uploaded on the site that are not searched by the clinicaltrials.gov search engine.

Our searches identified a total of 105 trials conducted or planned under EFIC. As of April 2022, 77 trials have been completed, 10 are recruiting, 10 have been registered on clinicaltrials.gov but are not yet active, and 5 were planned, including documented community engagement activities, but were not done for a variety of reasons. We also identified 3 trials that are in early planning stages (but are not yet registered). Of 77 completed trials, 9 (11.7%) were identified as pilot or feasibility trials. Pilot studies had on average significantly fewer subjects enrolled (mean=369.2) than full trials (mean=1240.7) (z=2.045, p=0.041 by nonparametric Wilcoxon rank-sum test). Eleven of the trials involved children, including 6 of the completed trials. Notably, the first pediatric trial first enrolled subjects in 2007, reflecting a lag in use of EFIC for trials involving children.^14^ Table 1 presents the medical conditions addressed by the trials, distinguishing between adult and pediatric studies. As shown by others, cardiac arrest and injuries caused by trauma were the focus of over 2/3 of these trials.

**Table 1.** Health condition/disease focus of trial

| **Condition** | **Adult Trials**<br>**<u>N (%)</u>** | **Pediatric**<br>**<u>Trials N(%)</u>** |
| --- | --- | --- |
| Cardiac arrest | 28 (29.5) | 2 (20.0) |
| TBI | 13 (13.7) | 1 (10.0) |
| Other trauma* | 28 (29.5) | 2 (20.0) |
| Respiratory failure | 9 (9.5) | 1 (10.0) |
| Stroke | 8 (8.4) | 0 |
| Other** | 8 (8.4) | 4 (40.0) |
| Unknown | 1 (1.0) | 0 |
| Totals | 95 | 10 |

\* Includes hemorrhage and shock.
\*\* Includes sepsis, seizure, agitation, burns, hemorrhage secondary to surgery,
Psychostimulant Drug-Induced Toxicity, and critical illness not otherwise specified.

To examine whether the number of trials is changing over time, we grouped trials by the year in which recruitment began into 5-year periods. As shown in Table 2, after a run-in period in the first several years after the EFIC rule was adopted, the number of trials initiated in each period has been remarkably stable, with a small drop in the last years possibly attributable to the Covid-19 pandemic. Note that two EFIC trials were begun in 1994, and each was modified to use an EFIC waiver in 1996.^20-22^

**Table 2.** Number of trials initiating enrollment in 5-year periods since the EFIC rule was adopted (through April 1, 2022)

| Enrollment |  | Trial Status |  |  |  |  |
| --- | --- | --- | --- | --- | --- | --- |
| <u>Began</u> |  | <u>Complete</u> | <u>Ongoing</u> | <u>Registered</u> | <u>Planned</u> | <u>Total</u> |
| 1996-2001 |  | 7 | 0 | 0 | 0 | 7 |
| 2002-2006 |  | 18 | 0 | 0 | 0 | 18 |
| 2007-2011 |  | 20 | 0 | 0 | 0 | 20 |
| 2012-2016 |  | 18 | 1 | 0 | 0 | 19 |
| 2017-2021 |  | 10 | 4 | 0 | 0 | 14 |
| Dates unknown |  | 4 | 5 | 0 | 0 | 9 |
| Not yet recruiting |  | 0 | 0 | 10 | 0 | 10 |
| Abandoned |  | 0 | 0 | 0 | 5 | 5 |
| <u>Planned</u> |  | <u>0</u> | <u>0</u> | <u>0</u> | <u>3</u> | <u>3</u> |
| Totals |  | 77 | 10 | 10 | 8 | 105 |

Like Klein et al.^7^ and Feldman et al.,^8^ we examined early termination of trials. As shown in Table 3, nearly half of full trials, 30 of 68 (44.1%), were halted early. More than half (17/30; note that 2 trials were terminated for both futility and safety concerns) of these terminations were attributed to futility and safety issues.

**Table 3.** Early termination and reasons given by authors, for full trials and pilot/feasibility trials

| **Reasons for**<br>**Termination \*** | **Full trials**<br>**N (68)** | **Pilot trials**<br>**N (9)** |
| --- | --- | --- |
| Not terminated | 38 | 6 |
| Futility | 15 | 0 |
| Low recruitment | 9 | 1 |
| Safety issues | 4 | 0 |
| Superiority | 1 | 0 |
| Funding issues | 4 | 1 |
| Covid-19 | 2 | 2 |
| Other reasons | 2 | 0 |

\* Totals are superadditive, because eight trials gave multiple reasons for termination.

## DISCUSSION

The main purpose of this study has been to update earlier reviews, which reports appeared to yield slightly different samples of trials, and to provide the census of trials to the research community for further examination. One main finding is that, of 77 completed trials for which we have recruitment dates, 63 were complete by the end of 2016. This suggests that the searches conducted by Klein et al.,^7^ Feldman et al.,^8^ and Haggins et al.^10^ were incomplete, even though these authors were using the FDA’s public repository. This raises an obvious question about why the repository appears to be incomplete, about which we have no insights. The fact that the FDA is highly protective of proprietary information (such as Investigative New Drug (IND) and Investigative Device Exemption (IDE) numbers and related information) makes it impossible for external researchers to identify EFIC trials or cross-check search results.

This finding points as well to the difficulty of searching for and identifying these trials. We find this to be somewhat ironic, given that the EFIC rule is predicated upon public disclosures and community engagement, even requiring “Public disclosure of sufficient information following completion of the clinical investigation to apprise the community and researchers of the study, including the demographic characteristics of the research population, and its results.”^23^ Simply, it should be much easier for researchers and others to identify and examine these trials. While many researchers we reached out to have been open and helpful, roughly a third were not (and one was outright hostile).

Second, we find that the number of trials conducted over time has been remarkably constant. Many of the trials are run under the auspices of trial consortia, including the Resuscitation Outcomes Consortium,^24^ the Neurological Emergencies Treatment Trials (NETT) Network,^25^ the Strategies to Innovate EmeRgENcy Care Clinical Trials Network (SIREN),^26^ the Linking Investigations in Trauma and Emergency Services (LITES) Network,^27^ the Pediatric Emergency Care Applied Research Network (PECARN),^28^ the Trans Agency Consortium for Trauma-Induced Coagulopathy (TACTIC),^29^ and the Trial Innovation Network,^30^ among others. These groups have developed the capacity and methods for implementing EFIC trials, and in some cases appear to have maintained stable portfolios of trials amongst network collaborators.

Third, we find that roughly 45% of the full (not pilot or feasibility) EFIC trials that have been completed were closed to recruitment early. This rate of termination is roughly the same as reported by Klein et al.^7^ (42%) and higher than that reported by Feldman et al.^8^ (at least 24%). This rate appears higher than for clinical trials in general. For example, Williams and colleagues found that 12% of 7646 clinical trials registered on clinicaltrials.gov with results posted prior to February 2013 were stopped early, predominantly because of enrollment difficulties.^31^ Poor accrual was also found to be the predominant reason for early termination in a study reporting that 10.9% of 6279 cardiovascular clinical trials registered on clinicaltrials.gov between February 2000 and January 2013 were terminated early,^32^ and an overlapping study that found 11% of 8900 completed cardiovascular trials on clinicaltrials.gov that started between January 2006 and December 2015 stopped early.^33^ Similar results have been seen in cancer research, with one study finding that 20% of 7776 phase II-III adult cancer trials failed to complete,^34^ and another reporting early termination of 19.8% of 886 clinical trials in glioblastoma multiforme completed between 2003 and 2020.^35^

Only one other review we have found has shown a similarly high rate of early termination. Strong and colleagues reported that 41% of 96 trials for treatment of acute stroke, having target enrollments of more than 100 patients and appearing in 9 leading clinical journals between 2013 and 2020, were terminated early, predominantly due to benefit (10.4%), logistical issues (10.4%), and futility (8.3%), with 1 trial stopped for safety.^36^ In contrast, only 1 EFIC trial was terminated early for superiority, the ARREST trial,^37^ while 17 full trials (25.0%) were halted due at least in part to futility or safety (Table 3). This perhaps reflects the difficulty of improving emergency care, but it also suggests that the EFIC trials, as a class, are different from other kinds of clinical research.

The high rate of early termination for futility and safety suggests that the balancing of the potential benefits of developing interventions and designing studies for patients facing life-threatening conditions for which existing therapies are “unproven or unsatisfactory,”^38^ against the risks of experimental interventions – including the likelihood that they will be ineffective or dangerous – is different than for other kinds of research. While greater risk may be acceptable in a domain where the potential gains from therapeutic advances can have direct life-saving effects, continued monitoring of EFIC trial performance and outcomes is critical to understanding whether the balance achieved in practice is appropriate. For example, one ethical requirement for randomized clinial trials is equipoise, the condition of community uncertainty about which of two (or more) treatments being compared is better.^3,39^ Examination of trial outcomes may help refine approaches to understanding equipoise, with the goal of improving *ex-ante* decisions about proposed trials by researchers, Institutional Review Boards, and the FDA.

The main limitation of this study is our inability to state our confidence in our final census of trials. We have triangulated sources, drawing on review articles that examine EFIC trials as well as related community outreach and other topics, contacting known researchers, and searching broadly for secondary referrals to trials. Some uncertainty (identified in the Supplement) remains due to ambiguous language in published papers. Finally, we posted a draft of this manuscript and the supplemental list of trials on the medrxiv preprint server, seeking input and additional information from the research community about the list.^40^

In conclusion, we have developed a census of trials planned or conducted under the US EFIC regulations. The list of trials is available to others in the Supplement, as we continue our own analyses of these trials. We believe the continued in-depth collection, analysis, and dissemination of details of these trials is warranted.

## Data Availability

The list of trials with sources (publications, NCT numbers) is provided in the Supplement.

## Acknowledgements

This paper was prepared in partial fulfillment of requirements for the Masters of Bioethics degree (KS). JFM conceived of the idea for the study, KS and JFM developed the approach and coding scheme, searched for trials, and read and coded papers and protocols. JFM contacted authors and collected responses, and performed the quantitative analysis. KS wrote the first version of the paper, and JFM managed subsequent editing. The authors contributed equally to the coding, checking, and interpretation of data, and the editing of the manuscript. The authors thank Jason McMullan and other study investigators and review authors who responded to our inquiries and provided information about emergency trials, and Will Feldman, JFM’s summer 2022 research ethics class, and several anonymous reviewers for comments. Responsibility for the study is solely that of the authors. Funding: none.

