## Supplement for "A Census of Clinical Trials Conducted Under the US Exception from Informed Consent Rule"

Krista L. Snyder & Jon F. Merz. A Census of Clinical Trials Conducted Under the US Exception from Informed Consent. Supplement: List of completed, ongoing, planned but not yet enrolling, abandoned, and early-stage planning trials, as of April, 2022.

| **Main results paper or other trial identification** | **Acronym** | **Notes** |
| --- | --- | --- |
| **Completed** |  |  |
| Abella BS, Edelson DP, Kim S, Retzer E, Myklebust H, Barry AM, et al. CPR quality improvement during in-hospital cardiac arrest using a real-time audiovisual feedback system. Resuscitation 2007; 73:54-61. | none | IDE G020121; NCT00228293; compared intervention group to historic controls (not an RCT). |
| Adelson PD, Wisniewski SR, Beca J, et al. Comparison of hypothermia and normothermia after severe traumatic brain injury in children (Cool Kids): a phase 3 randomised controlled trial. Lancet Neurol 2013; 12:546-553. | Cool Kids | NCT00222742; 5 of 15 sites secured (in US) an EFIC waiver |
| Alldredge BK, Gelb AM, Isaacs SM, et al. A comparison of lorazepam, diazepam, and placebo for the treatment of out-of-hospital status epilepticus. N Engl J Med 2001;345: 631-637. | PHTSE | Protocol (details of EFIC implementation) published at: Lowenstein DH, Alldredge BK, Allen F, et al. The pre-hospital treatment of status epilepticus (PHTSE) study: Design and methodology. Control Clin Trials 2001; 22: 290-309. |
| Aufderheide TP, Frascone RJ, Wayne MA, et al. Standard cardiopulmonary resuscitation versus active compression-decompression cardiopulmonary resuscitation with augmentation of negative intrathoracic pressure for out-of-hospital cardiac arrest: a randomised trial. Lancet 2011; 377:301-311. | ResQ Trial | IDE G050062; NCT00189423; the link in the Lancet article to the protocol doesn't work, and it takes you to the website of Zoll; community outreach described in: Salzman JG, Frascone RJ, Godding BK, Provo TA, Gertner E. Implementing emergency research requiring exception from informed consent, community consultation, and public disclosure. Ann Emerg Med 2007; 50:448-455. |
| Aufderheide TP, Nichol G, Rea TD, et al. A trial of an impedance threshold device in out-of-hospital cardiac arrest. N Engl J Med 2011; 365:798-806. | ROC PRIMED | NCT00394706 |
| Aufderheide TP, Pirrallo RG, Provo TA, Lurie KG. Clinical evaluation of an inspiratory impedance threshold device during standard cardiopulmonary resuscitation in patients with out-of-hospital cardiac arrest. Crit Care Med 2005; 33:734-740. | none | IDE 980125 |
| Aufderheide TP, Sigurdsson G, Pirrallo RG, et al. Hyperventilation-induced hypotension during cardiopulmonary resuscitation. Circulation 2004; 109:1960-1965. | none | This was an observational study of some of the subjects in the Crit Care Med 2005 trial; source is that paper. |
| Balamuth F, Kittick M, McBride P, et al. Pragmatic pediatric trial of balanced versus normal saline fluid in sepsis: the PRoMPT BOLUS randomized controlled trial pilot feasibility study. Acad Emerg Med 2019; 26:1346-1356. | PRoMPT BOLUS | IND 13698; NCT03340805 |
| Bulger EM, Jurkovich GJ, Nathens AV, et al. Hyptertonic resuscitation of hypovolemic shock after blunt trauma: a randomized controlled trial. Arch Surg 2008; 143:139-148. | none | IND 10292; NCT00113685 |
| Bulger EM, May S, Brasel KJ, et al. Out-of-hospital resuscitation following severe traumatic brain injury: a randomized controlled trial. JAMA 2010; 304:1455-1464. | ROC-HS/TBI | IND 12505; NCT00316004 |
| Bulger EM, May S, Kerby JD, et al. Out-of-hospital hypertonic resuscitation after traumatic hypovolemic shock: a randomized, placebo controlled trial. Ann Surg 2011; 253:431-441. | ROC-HS/S | IND 12506; NCT00316017; community consultation described in: Nelson M, Schmidt TA, DeIorio NM, McConnell KJ, Griffiths DE, McClure KB. Community consultation methods in a study using exception to informed consent. Prehosp Emerg Care 2008; 12:417-425. |
| Callaway CW, Hostler D, Doshi AA, et al. Usefulness of vasopressin administered with epinephrine during out-of-hospital cardiac arrest. Am J Cardiol 2006; 98:1316-1321. | none | IND 66463 |
| Carroll TG, Dimas VV, Raymond TT. Vasopressin rescue for in-pediatric intensive care unit cardiopulmonary arrest refractory to initial epinephrine dosing: a prospective feasibility pilot trial. Pediatr Crit Care Med 2012; 13:265-272. | none | IND 77955; NCT 00628550; community outreach described in: Raymond TT, Carroll TG, Sales G, Morris MC. Effectiveness of the informed consent process for a pediatric resuscitation trial. Pediatrics 2010; 125:e866-e875. |
| Chamberlain JM, Kapur J, Shinnar S, et al. Efficacy of levetiracetam, fosphenytoin, and valproate for established status epilepticus by age group (ESETT): a double-blind, responsive-adaptive, randomised controlled trial. Lancet 2020; 395:1217-1224. | ESETT | IND 119756; NCT01960075; trial terminated early for futility, but then extended enrollment in children to compare outcomes. See Kapur J, Elm J, Chamberlain JM, et al. Randomized trial of three anticonvulsant medications for status epilepticus. New Engl J Med 2019; 381:2103-2113. |
| Chamberlain JM, Okada P, Holsti M, et al. Lorazepam vs diazepam for pediatric status epilepticus: a randomized clinical trial. JAMA 2014; 311:1652-1660. | STATUS2 | IND 79010; NCT00621478; community consultation described in Holsti M, Zemek R, Baren J, et al. Variation of community consultation and public disclosure for a pediatric multi-centered "exception from informed consent" trial. Clin Trials 2015; 12:67-76. Note that an earlier pharmacodynamic study of Lorazepam was done by these investigators, with consent: NCT00114569. |
| Clifton GL, Miller ER, Choi SC, et al. Lack of effect of induction of hypothermia after acute brain injury. New Engl J Med 2001; 344:556-563. | NABISH | Note: this trial started with consent and was halted because of poor enrollment; it was granted a complete waiver by the Secretary of DHHS, but actually applied the EFIC standards, including public notification and securing surrogate consent when possible (Clifton, personal communication, 2021). See: Clifton GL, Knudson P, McDonald M. Waiver of consent in studies of acute brain injury. J Neurotrauma 2002; 19:1121-1126. |
| Clifton GL, Valadka A, Zygun D, et al. Very early hypothermia induction in patients with severe brain injury (the National Acute Brain Injury Study: Hypothermia II): a randomised trial. Lancet Neurol 2011; 10:131-139. | NABISH-II | IDE G080088; NCT00178711; community consultation in Jackson, MS, described in: Dix ES, Esposito D, Spinosa F, Olson N, Chapman S. Implementation of community consultation for waiver of informed consent in emergency research: one institutional review board's experience. J Investig Med 2004; 52:113-116 (note: no subjects were enrolled in Jackson, attributable to the warm IDE G960214; IDE G960214; IDE G960214; IDE G960214; IDE G960214; environment). |
| Clinical Investigation of the VEST-CPR System in Adults; IDE G960214 | VEST-CPR | Community consultation described in: Kremers MS, Whisnant DR, Lowder LS, Gregg L. Initial experience using the food and drug administration guidelines for emergency research without consent. Ann Emerg Med 1999; 33:224-229. See also Shah AN, Sugarman J. Protecting research subjects under the waiver of informed consent for emergency research: experiences with efforts to inform the community. Ann Emerg Med 2003; 41:72-78. |
| Cohn SM, McCarthy J, Stewart RM, Jonas RB, Dent DL, Michalek JE. The impact of low-dose vasopressin on trauma outcome: prospective randomized study. World J Surg 2011; 35:430-439. | none | IND 74154; NCT00420407 |
| Cotton BA, Podbielski J, Camp E, et al. A randomized controlled pilot trial of modified whole blood versus component therapy in severely injured patients requiring large volume transfusions. Ann Surg 2013; 258:527-533. | none | NCT01227005; see: Erratum, Ann Surg 2013; 260:178 (changing the reference from the EFIC rule to OPRR Reports 97-01, which suggests the trial did not comply with all of the FDA requirements). |
| Driver BE, Prekker ME, Moore JC, Schick AL, Reardon RF, Miner JR. Direct versus video laryngoscopy using the C-MAC for tracheal intubation in the emergency department, a randomized controlled trial. Acad Emerg Med 2016; 23:433-439. | none | NCT01710891 |
| Dutton RP, Mackenzie CF, Scalea TM. Hypotensive resuscitation during active hemorrhage: impact on in-hospital mortality. J Trauma 2002; 52:1141-1146. | Fluid Resuscitation in Trauma (FRT) |  |
| Gonzalez E, Moore EE, Moore HB, et al. Goal-directed hemostatic resuscitation of trauma-induced coagulopathy: a pragmatic randomized clinical trial comparing a viscoelastic assay to conventional coagulation assays. Ann Surg 2016; 263:1051-1059. | none |  |
| Guyette FX, Brown JB, Zenati MS, et al. Tranexamic acid during prehospital transport in patients at risk for hemorrhage after injury: a double-blind, placebo-controlled, randomized clinical trial. JAMA Surg 2021; 156:11-20. | STAAMP | NCT02086500 |
| Hallstrom A, Rea TD, Sayre MR, et al. Manual chest compressions vs use of an automated chest compression device during resuscitation following out-of-hospital cardiac arrest: a randomized trial. JAMA 2006; 295:2620-2628. | ASPIRE | NCT00120965; secondary analysis of data appears in: Paradis NA, Young G, Lemeshow S, Brewer JE, Halperin HR. Inhomogeneity and temporal effects in AutoPulse Assisted Prehospital International Resuscitation–an exception from consent trial terminated early. Am J Emerg Med 2010; 28:391-398. |
| Hergenroeder GW, Yokobori S, Choi HA, et al. Hypothermia for patients requiring evacuation of subdural hematoma: a multicenter randomized clinical trial. Neurocrit Care 2022; 36:560-572. | HOPES | NCT02064959 |
| Holcomb JB, Tilley BC, Baraniuk S, et al. Transfusion of plasma, platelets, and red blood cells in a 1:1:1 vs a 1:1:2 ratio and mortality in patients with severe trauma: the PROPPR randomized clinical trial. JAMA 2015; 313:471-482. | PROPPR | IND 14929; NCT01545232; community outreach in Toronto described in: Henry B, Perez A, Trpcic S, Rizoli S, Nascimento B. Protecting study participants in emergency research: is community consultation before trial commencement enough? Trauma Surg Acute Care Open 2017; 2:e000084. |
| Hsu CH, Meurer WJ, Domeier R, et al. Extracorporeal cardiopulmonary resuscitation for refractory out-of-hospital cardiac arrest (EROCA): results of a randomized feasibility trial of expedited out-of-hospital transport. Ann Emerg Med 2021; 78:92-101. | EROCA | NCT03065647 |
| HU23F2G for Hemorrhagic Shock; IND 7371 | none | Trial described in Feldman et al. 2018 |
| Jacoby J, Hellter M, Nicholas J, et al. Etomidate versus midazolam for out-of-hospital intubation: a prospective, randomized trial. Ann Emerg Med 2006; 47:525-530. | none |  |
| Kern KB, Radsel P, Jentzer JC, et al. Randomized pilot clinical trial of early coronary angiography versus no early coronary angiography after cardiac arrest without ST-segment elevation: the PEARL study. Circulation 2020; 142:2002-2012. | PEARL | NCT02387398; public engagement described in: Eubank L, Lee KS, Seder DB, et al. Approaches to community consultation in exception from informed consent: analysis of scope, efficiency, and cost at two centers. Resuscitation 2018; 130:81-87. |
| Kidwell CS, Jahan R, Gornbein J, et al. A trial of imaging selection and endovascular treatment for ischemic stroke. N Engl J Med 2013; 368:914-923. | MR RESCUE | IDE G050077; NCT00389467; protocol in Kidwell CS, Jahan R, Alger JR, et al. Design and rationale of the Mechanical Retrieval and Recanalization of Stroke Clots Using Embolectomy (MR RESCUE) trial. Intern J Stroke 2014; 9:110-116. |
| Kim F, Maynard C, Dezfulian C, et al. Effect of out-of-hospital sodium nitrite on survival to hospital admission after cardiac arrest: a randomized clinical trial. JAMA 2021; 325:138-145. | SNOCAT | NCT03452917 |
| Kim F, Nichol G, Maynard C, et al. Effect of prehospital induction of mild hypothermia on survival and neurological status among adults with cardiac arrest: a randomized clinical trial. JAMA 2014; 311:45-52. | none | IND 69382; NCT00391469 |
| Kim F, Olsufka M, Maynard C, et al. Pilot randomized clinical trial of prehospital induction of mild hypothermia in out-of-hospital cardiac arrest patients with a rapid infusion of 4°C normal saline. Circulation 2007; 115:3064-3070. | none | NCT00329563 |
| Kudenchuck PJ, Brown SP, Daya M, et al. Amiodarone, lidocaine, or placebo in out-of-hospital cardiac arrest. N Engl J Med 2016; 374:1711-1722. | ROC-ALPS | IND 110280; NCT01401647 |
| Linakis SW, Kuppermann N, Stanley RM, et al. Enrollment with and without exception from informed consent in a pilot trial of tranexamic acid in children with hemorrhagic injuries. Acad Emerg Med 2021; 28:1421-1429. | TIC-TOC | NCT04387305 for full trial; community engagement described in: Powers PE, Shore KK, Perez S, et al. Public deliberation as a novel method for an exception from informed consent community consultation. Acad Emerg Med 2019; 26:1158-1168. |
| Longstreth WT Jr, Fahrenbruch CE, Olsufka M, Walsh TR, Copass MK, Cobb LA. Randomized clinical trial of magnesium, diazepam, or both after out-of-hospital cardiac arrest. Neurology 2002; 59:506-514. | none | IND 52523 |
| Martel M, Sterzinger A, Miner J, Clinton J, Biros M. Management of acute undifferentiated agitation in the emergency department: a randomized double-blind trial of Droperidol, Ziprasidone, and Midazolam. Acad Emerg Med 2005; 12:1167-1172. | none | “Study A" |
| Martel ML, Driver BE, Miner JR, Biros MH, Cole JB. Randomized double-blind trial of intramuscular Droperidol, Ziprasidone, and Lorazepam for acute undifferentiated agitation in the emergency department. Acad Emerg Med 2021; 28:421-434. | none | "Study B"; publication delayed until 2021. |
| Matchett G, Gasanova I, Riccio CA, et al. Etomidate versus ketamine for emergency endotracheal intubation: a randomized clinical trial. Intensive Care Med 2022; 48:78-91; doi:10.1007/s00134-021-06577-x | EvK Clinical Trial | NCT02643381 |
| Moore EE, Moore FA, Fabian TC, et al. Human polymerized hemoglobin for the treatment of hemorrhagic shock when blood is unavailable: the USA multicenter trial. J Am Coll Surg 2009; 208:1-13. | USA Multicenter Trial | IND 10719; NCT00076648; public engagement in San Antonio described in: Longfield JN, Morris MJ, Moran KA, Kragh JF Jr, Wolf R, Baskin TW. Community meetings for emergency research community consultation. Crit Care Med 2008; 36:731-736. |
| Moore HB, Moore EE, Chapman MP, et al. Plasma-first resuscitation to treat haemorrhagic shock during emergency ground transportation in an urban area: a randomised trial. Lancet 2018; 392:283-291. | COMBAT | IND 15216; NCT01838863; community consultation described in: Chin TL, Moore EE, Coors ME, et al. Exploring ethical conflicts in emergency trauma research: the COMBAT (Control of Major Bleeding After Trauma) study experience. Surgery 2015; 157:10-19. |
| NCT00401882; IND 74308; Treatment of Ventricular Tachyarrhythmias Refractory To Shock With Beta Blockers: The SHOCK and BLOCK Trial | Shock & Block | Unpublished |
| NCT00805818 & NCT01366820; IND 111724; Study of NNZ-2566 in Patients With Traumatic Brain Injury (INTREPID2566) | INTREPID | Unpublished |
| NCT00973674; Resuscitative Endocrinology:Single-dose Clinical Uses for Estrogen-Traumatic Brain Injury | RESCUE-TBI | Trial was run parallel to Wigginton et al., NCT00973102 |
| NCT01325870; IDE G080169; RESQPRO phase I trial | RESQPRO |  |
| IND 122444; NCT01823328; Ketamine Versus Etomidate for Rapid Sequence Intubation | none |  |
| NCT03079102; Inhaled Nitric Oxide After Out-of-Hospital Cardiac Arrest (iNOOHCA) | iNOOHCA |  |
| NCT03119571; ACCESS to the Cardiac Cath Lab in Patients Without STEMI Resuscitated From Out-of-hospital VT/VF Cardiac Arrest | ACCESS | community engagement described in: Hsu CH, Fowler J, Cranford JA, Thomas MP, Neumar RW. Integration of social media with targeted emails and in-person outreach for exception from informed consent community consultation. Acad Emerg Med 2021; 29:217-227; doi:10.1111/acem.14377. |
| NCT03189433; Efficacy and Safety Study of Ryanodex as Adjuvant Treatment in Subjects With Psychostimulant Drug-Induced Toxicity (PDIT) | none |  |
| NCT03763929; Efficacy and Safety of Trans Sodium Crocetinate (TSC) for Treatment of Suspected Stroke | PHAST-TSC |  |
| Nichol G, Leroux B, Wang H, et al. Trial of continuous or interrupted chest compressions during CPR. N Engl J Med 2015; 373:2203-2214. | ROC Continuous Chest Compressions Trial | NCT01372748 |
| O'Malley et al. cricoid versus bimanual laryngoscopy study | none | Community engagement described in: O'Malley GF, Giraldo P, Deitch K, et al. A novel emergency department-based community notification method for clinical research without consent. Acad Emerg Med 2017; 24:721-731 (includes limited information about enrollment in the underlying trial). |
| Pusateri AE, Le TD, Keyloun JW, et al. Early abnormal fibrinolysis and mortality in patients with thermal injury: a prospective cohort study. BJS Open 2021; zrab017. doi:10.1093/bjsopen/zrab017 | none | Prospective cohort study. |
| Reynolds PS, Michael MJ, Cochran ED, Wegelin JA, Spiess BD. Prehospital use of plasma in traumatic hemorrhage (The PUPTH Trial): study protocol for a randomised controlled trial. Trials 2015; 16:321. | PUPTH | IND 15910; NCT02303964 |
| Robertson CS, Hannay HJ, Yamal JM, et al. Effect of erythropoietin and transfusion threshold on neurological recovery after traumatic brain injury: a randomized clinical trial. JAMA 2014; 312:36-47. | none | IND 100681; NCT00313716; recruitment data analyzed in Yamal JM, Robertson CS, Rubin ML, Benoit JS, Hannay HJ, Tilley BC. Enrollment of racially/ethnically diverse participants in traumatic brain injury trials: effect of availability of exception from informed consent. Clin Trials 2014; 11:187-194. |
| Rowell SE, Meier EN, McKnight B, et al. Effect of out-of-hospital tranexamic acid vs placebo on 6-month functional neurologic outcomes in patients with moderate or sever traumatic brain injury. JAMA 2020; 324:961-974. | Prehospital TXA for TBI Trial | IND 119858; NCT01990768 |
| Saver JL, Starkman S, Eckstein M, et al. Prehospital use of magnesium sulfate as neuroprotection in acute stroke. N Engl J Med 2015; 372:528-536. | FAST-MAG | NCT00059332 |
| Schreiber MA, Meier EN, Tisherman SA, et al. A controlled resuscitation strategy is feasible and safe in hypotensive trauma patients: results of a prospective randomized pilot trial. J Trauma Acute Care Surg 2015; 78:687-697. | HypoResus | IND 110699; NCT01411852 |
| Selker HP, Beshansky JR, Sheehan PR, et al. Out-of-hospital administration of intravenous glucose-insulin-potassium in patients with suspected acute coronary syndromes: the IMMEDIATE randomized controlled trial. JAMA 2012; 307:1925-1933. | IMMEDIATE | IND 70376; NCT00091507; community outreach described in: Beshansky JR, Sheehan PR, Klima KJ, Hadar N, Vickery EM, Selker HP. A community consultation survey to evaluate support for and success of the IMMEDIATE trial. Clin Trials 2014; 11:178-186. |
| Silbergleit R, Durkalski V, Lowenstein D, et al. Intramuscular versus intravenous therapy for prehospital status epilepticus. N Engl J Med 2012; 366:591-600. | RAMPART | IND 102254; NCT00809146; community consultation described in: Dickert NW, Govindarajan P, Harney D, et al. Community consultation for prehospital research: experiences of study coordinators and principal investigators. Prehosp Emerg Care 2014;18:274-281; Govindarajan P, Dickert NW, Meeker M, et al. Emergency research: using exception from informed consent, evaluation of community consultations. Acad Emerg Med 2013; 20:98-103; Vohra T, Chebl RB, Miller J, Russman A, Baker A, Lewandowski C. Improving community understanding of medical research: audience response technology for community consultation for exception to informed consent. West J Emerg Med 2014; 15:414-418; Biros MH, Sargent C, Miller K. Community attitudes towards emergency research and exception from informed consent. Resuscitation 2009; 80:1382-1387. |
| Sims CA, Holena D, Kim P, et al. Effect of low-dose supplementation of arginine vasopressin on need for blood product transfusions in patients with trauma and hemorrhagic shock: a randomized clinical trial. JAMA Surg 2019; 154:994-1003. | AVERTshock | IND 110314; NCT01611935; community engagement described in: Sims CA, Isserman JA, Holena D, et al. Exception from informed consent for emergency research: consulting the trauma community. J Trauma Acute Care Surg. 2013; 74:157-165. |
| Sloan EP, Koenigsberg M, Gens D, et al. Diaspirin cross-linked hemoglobin (DCLHb) in the treatment of severe traumatic hemorrhagic shock: a randomized controlled efficacy trial. JAMA 1999; 282:1857-1864. | DCLHb Traumatic Hemorrhagic Shock Study | Baxter's Hemassist Phase III trial, INDA 6859; documents collected at: https://www.circare.org/subjects/dclhb.htm; public engagement described in Santora TA, Cowell V, Trooskin SZ. Working through the public disclosure process mandated by use of 21 CFR 50.24 (exception to informed consent): guidelines for success. J Trauma 1998; 45:907-913. See http://web.archive.org/web/19990125085353/http://dclhb.er.uic.edu/ |
| Smischney NJ, Nicholson WT, Brown DR, et al. Ketamine/propofol admixture vs etomidate for intubation in the critically ill: KEEP PACE randomized clinical trial. J Trauma Acute Care Surg 2019; 87:883-891. | KEEP PACE | NCT02105415; protocol: Smischney NJ, Hoskote SS, de Moraes AG, et al. Ketamine/propofol admixture (ketofol) at induction in the critically ill against etomidate (KEEP PACE trial): study protocol for a randomized controlled trial. Trials 2015; 16:177. |
| Smith WS, Sung G, Saver JL, et al. Mechanical thrombectomy for acute ischemic stroke: final results of the Multi MERCI trial. Stroke 2008; 39:1205-1212. | Multi-MERCI | IDE G020163; nonrandomized, single-arm multi-center trial - 15 sites, 2 in Canada; paper alludes to waiver at some sites, but unclear if this actually happened. |
| Smith WS, Sung G, Starkman S, et al. Safety and efficacy of mechanical embolectomy in acute ischemic stroke: results of the MERCI trial. Stroke 2005; 36:1432-1440. | MERCI | Nonrandomized, single-arm multi-center trial - 25 US sites, 2 secured EFIC waivers. |
| Sperry JL, Guyette FX, Brown JB, et al. Prehospital plasma during air medical transport in trauma patients at risk for hemorrhagic shock. New Engl J Med 2018; 379:315-326. | PAMPer | NCT01818427 |
| Sperry JL. Pragmatic Prehospital Group O Whole Blood Early Resuscitation Trial (PPOWER) | PPOWER | NCT03477006 |
| Spinella PC, Thomas KA, Turnbull IR, et al. The immunologic effect of early intravenous two and four gram bolus dosing of tranexamic acid compared to placebo in patients with severe traumatic bleeding (TAMPITI): a randomized, double-blind, placebo-controlled, single-center trial. Front Immunol 2020; 11(2085):1-16. | TAMPITI | NCT02535949 |
| Stiell IG, Nichol G, Leroux BG, et al. Early versus later rhythm analysis in patients with out-of-hospital cardiac arrest. N Engl J Med 2011; 365:787-797. | ROC PRIMED | NCT00394706 (same as Aufderheide et al. 2011) |
| The Public Access Defibrillation Trial Investigators. Public-access defibrillation and survival after out-of-hospital cardiac arrest. N Engl J Med 2004; 351:637-646. | PAD Trial | IDE G980067; protocol published at: Ornato JP, McBurnie MA, Nichol G, et al. The Public Access Defibrillation (PAD) Trial: study design and rationale. Resuscitation 2003; 56:135-147; community consultation described in: Mosesso VN, Brown LH, Greene HL, et al. Conducting research using the emergency exception from informed consent: the public access defibrillation (PAD) trial experience. Resuscitation 2004; 61:29-36. |
| Wang HE, Schmicker RH, Daya MR, et al. Effect of a strategy of initial laryngeal tube insertion vs endotracheal intubation on 72-hour survival in adults with out-of-hospital cardiac arrest: a randomized clinical trial. JAMA 2018; 320:769-778. | PART | NCT02419573; protocol: Wang HE, Prince DK, Stephens SW, et al. Design and implementation of the Resuscitation Outcomes Consortium Pragmatic Airway Resuscitation Trial (PART). Resuscitation 2016; 101:57-64. |
| Wigginton JG, Pepe PE, Warren V, et al. Feasibility and experience of using exception from informed consent in a pilot study of immediate estrogen infusion for hypotensive trauma patients. Crit Care 2013; 17(Suppl 2):P288. | RESCUE Shock | NCT00973102 |
| Wik L, Olsen JA, Persse D, et al. Manual vs. integrated automatic load-distributing band CPR with equal survival after out of hospital cardiac arrest: the randomized CIRC trial. Resuscitation 2014; 85:741-748. | CIRC | NCT00597207; sponsored by Zoll Medical; protocol paper: Lerner EB, Persse D, Souders CM et al. Design of the Circulation Improving Resuscitation Care (CIRC) trial: a new state of the art design for out-of-hospital cardiac arrest research. Resuscitation 2011; 82:294-299. doi:10.1016/j.resuscitation.2010.11.013 |
| Wright DW, Yeatts SD, Silbergleit R, et al. Very early administration of progesterone for acute traumatic brain injury. N Engl J Med 2014; 371:2457-2466. | ProTECT III | IND 104188; NCT00822900 |
| Yannopoulos D, Bartos J, Raveendran G, et al. Advanced reperfusion strategies for patients with out-of-hospital cardiac arrest and refractory ventricular fibrillation (ARREST): a phase 2, single centre, open-label, randomised controlled trial. Lancet 2020; 396:1807-1816. | ARREST | NCT03880565 |
| **Currently recruiting (as of April 2022)** |  |  |
| Bernard F, Barsan W, Diaz-Arrastia R, Merck LH, Yeatts S, Shutter LA. Brain Oxygen Optimization in Severe Traumatic Brain Injury (BOOST-3): a multicentre, randomised, blinded-endpoint, comparative effectiveness study of brain tissue oxygen and intracranial pressure monitoring versus intracranial pressure alone. BMJ Open 2022; 12:e060188 | BOOST-3 | NCT03754114 |
| NCT02047028; Hyperbaric Oxygen Brain Injury Treatment Trial (HOBIT) | HOBIT | Protocol paper: Gajewski BJ, Berry SM, Barsan WG, et al. Hyperbaric oxygen brain injury treatment (HOBIT) trial: a multifactor design with response adaptive randomization and longitudinal modeling. Pharmaceutic Statist 2016; 15:396-404. |
| NCT03263117; SEdation Versus General Anesthesia for Endovascular Therapy in Acute Ischemic Stroke | SEGA | Inam ME, Sanzgiri A, Lekka E, et al. Exception from informed consent in the era of social media: The SEGA stroke trial experience. Brain Circ 2021; 7:253-258. |
| NCT03496883; Recombinant Factor VIIa (rFVIIa) for Hemorrhagic Stroke Trial | FASTEST | https://www.nihstrokenet.org/fastest/home |
| NCT03737786; SEACOAST 1- SEdAtion With COllAteral Support in Endovascular Therapy for Acute Ischemic Stroke | SEACOAST |  |
| NCT04100564; Prehospital Airway Control Trial | PACT |  |
| NCT04684719; Type O Whole Blood and Assessment of Age During Prehospital Resuscitation Trial | TOWAR |  |
| NCT04726410; Cold-stored Platelet Early Intervention in TBI | CriSP-TBI |  |
| Tisherman SA, Alam HB, Rhee PM, et al. Development of the emergency preservation and resuscitation for cardiac arrest from trauma clinical trial. Trauma Acute Care Surg 2017; 83:803-809; Tisherman SA. Emergency preservation and resusciation for cardiac arrest from trauma. Annals NY Acad Sci 2022; 1509:5-11. | EPR-CAT | NCT01042015; public notice provided at: https://www.umms.org/ummc/health-services/shock-trauma/news/body-cooling-study |
| Weiss SL, Balamuth F, Long E, et al. PRagMatic Pediatric Trial of Balanced vs nOrmaL Saline FlUid in Sepsis: study protocol for the PRoMPT BOLUS randomized interventional trial. Trials 2021; 22:776. | PRoMPT BOLUS | NCT04102371; this is the main trial following the pilot study in NCT03340805; website: https://www.research.chop.edu/prompt-bolus |
| **Not yet recruiting** |  |  |
| NCT03402035; Shock, Whole Blood, and Assessment of TBI S.W.A.T. (LITES TO 2) | SWAT |  |
| NCT03705286; Endotracheal Tubes to Prevent Ventilator-Associated Pneumonia | PreVent2 |  |
| NCT04019015; Prehospital Kcentra for Hemorrhagic Shock | KCENTRA |  |
| NCT04220619; RescueTEE for In-hospital Cardiac Arrest | ReTEECA trial |  |
| NCT04663087; Feasibility of Evaluating XSTAT Use in the Prehospital Setting (PhoXSTAT) | PhoXSTAT |  |
| NCT04667468; Cold Stored Platelet in Hemorrhagic Shock | CriSP-HS |  |
| NCT04704869; Early Use of Cryoprecipitate With Major Hemorrhage Protocol (MHP) Activation | none |  |
| NCT04849169; Investigation of the Ringer Perfusion Balloon Catheter | Ringer IDE |  |
| NCT05121324; Pediatric Dose Optimization for Seizures in Emergency Medical Services | PediDOSE |  |
| NCT05277896; Casey J, et al. Randomized Trial of Sedative Choice for Intubation (RSI) | RSI |  |
| **Abandoned** |  |  |
| NCT02191800; IDE G120223; A Randomized Comparative Multicenter, Open Label, Non- inferiority Study, to Compare the SolidAIRity Airway Stabilization System’s Ability to Prevent Unplanned Extubation Relative to Standard of Care in Critically Ill or Injured Subjects Requiring Emergency Department (ED) or Intensive Care Unit (ICU) Oral Intubation for Airway Management and Admission to the ICU3 | SolidAIRity | The trial was abandoned (note that a 510k license was granted, making the trial moot) |
| Nadkarni V (PI); Morris MC, Nadkarni VM, Ward FR, Nelson RM. Exception from informed consent for pediatric resuscitation research: community consultation for a trial of brain cooling after in-hospital cardiac arrest. Pediatrics 2004; 114:776-781. | THAPCA-IH | Underlying trial done with consent; NCT00880087; Molder FW, Silverstein FS, Holubkov R, et al. Therapeutic hypothermia after in-hospital cardiac arrest in children. New Engl J Med 2017; 376:318-329. |
| NCT02821364; Philadelphia Immediate Transport in Penetrating Trauma Trial | PIPT | Recruitment never opened |
| Robertson et al. L-arginine treatment of a reduced CBF after severe head injury | none | Trial funded, but not performed because stable xenon, which was necessary to study CBF, became unavailable in the US; community consultation described in: Contant C, McCullough LB, Mangus L, Robertson C, Valadka A, Brody B. Community consultation in emergency research. Crit Care Med 2006; 34:2049-2052. |
| Wang J (PI); Galbraith KL, Keck AS, Little C. Single-site community consultation for emergency research in a community hospital setting. Prehosp Emerg Care 2014; 18:328-334. | Cool Brain II | DOD funding was tentatively approved but did not come through. |
| **Planning stage (not yet registered as of April 2022)** |  |  |
| Bosson N, Hansen M, Gausche-Hill M, et al. Design of a novel clinical trial of prehospital pediatric airway management. Clin Trials 2022; 19:62-70. | Pedi-PART | not yet registered |
| Massive Transfusion in Children-2 (MATIC-2) Trial | MATIC-2 | not yet registered |
| TranExamic Atomized for Pediatric post-Operative Tonsillectomy hemorrhage (TEAPOT) | TEAPOT | not yet registered |
